# Booked Appointments System: An evaluation of the acceptability and feasibility of booked appointments in a large HIV clinic in Johannesburg, South Africa

**DOI:** 10.1101/2022.06.30.22277110

**Authors:** Lawrence Long, Rachel Batiancila, Sarah Girdwood, Pappie Majuba, Naomi Lince-Deroche

## Abstract

**Background:** Long queues and overcrowding are common in many of South Africa’s public healthcare facilities and may negatively impact on the quality of care provided. In HIV-related services, this problem may also affect retention. Healthcare facilities that rely on a system of patient appointments scheduled by day, and not by time, may exacerbate these issues. This study aims to improve retention and care by understanding the challenges and advantages related to the current system of booking appointments and assessing the potential for alternative systems.

**Methods:** The study was conducted in Johannesburg, South Africa at an outpatient HIV treatment clinic set within a large, urban secondary-level teaching hospital. The study is cross-sectional and includes structured interviews with providers and patients. Medical records are linked with patient interviews and observations to determine actual waiting times.

**Results:** 245 patients and 6 providers were interviewed. Of the patients interviewed, 64% were female, and 96% were Black African. Nearly a fifth of patients (19%) lost an average of USD 32 in income when attending their appointment. The most common reason for missing an appointment was that they could not be absent from work (40%), but despite this most respondents (65%) do not believe they face any challenges with the current system of booking appointments. Being able to arrive at a time convenient for them was considered a benefit by many (49%), but some (29%) recognized that this may result in overcrowding. The majority of respondents were in favor of appointments offered as a block of time in the morning or afternoon (88%) as well as appointments after work hours (85%). In comparison only just over half (58%) were in favor of booking appointments at a specific time. Most respondents (79%) believe the largest benefit to morning/afternoon block appointments would be the ability to show up at their own convenience during the block of time.

**Conclusions:** Patients value convenience highly. This may be explained by the need to be flexible around work schedules and transport options. It might be worth exploring a booking system in which patients are given a morning or afternoon appointment rather than a day. This may improve the distribution of patients throughout the day and as a result retention and care in a high-prevalence HIV setting.

## Introduction

Patient satisfaction has increasingly emerged as a meaningful parameter in assessing the quality of health care provision [1]. Waiting times are one factor by which patient satisfaction is measured within health care facilities. Despite existing research showing significant correlation between patient satisfaction and waiting times to see the doctor [2], public sector patients across Africa continue to experience long queues and wait times [3-6]. Across the continent, a number of studies have demonstrated that the major cause of dissatisfaction within public sector health care services is due to long waiting times [1].

The majority of South Africans (82.5%) are not registered on medical insurance schemes and are likely to receive all or part of their health services in the public sector [7]. In contrast though the public health sector expenditure in 2010 accounted for less than half of all healthcare expenditure in South Africa [8]. South Africa’s existing public health service delivery infrastructure has already been impacted by these constraints. Government health institutions have been found to be overcrowded with long waiting times and delays in service [9].

Long wait times can adversely impact patient health through complications linked to missed appointments, poor adherence to medication and delayed implementation of clinical programmes [10]. This plays a special role within HIV-endemic settings, such as South Africa. Studies have shown that long wait times among ART patients contribute to their dissatisfaction with clinic services, making them more likely to stop picking up their medication [11, 12]. This poses a major challenge to treatment adherence, where compliance among ART patients is paramount in reducing viral loads, preventing drug resistance, and increasing the chance of survival [13]. Furthermore, waiting in long queues has been found to exacerbate hunger; a unique barrier to adherence that ART patients face during the initial stages of treatment when the body requires extra nutrition and regains its strength [11, 12]. These patients have been found to favour ART programs where visits to the clinic are quicker and less frequent, which may alleviate both financial and emotional stress for patients who are most likely poor and in serious need of health services [14]. Health care providers must also carefully consider the cost implications of defaulting on treatment, as developing drug resistance can lead to more expensive second line ART treatment regimens [12, 15].

Waiting time satisfaction is an important component of overall patient satisfaction. This is recognised by the National Department of Health in the National Core Standards of 2011[16]. These standards outline a benchmark of quality care against which delivery of services can be monitored. Waiting times fall in the first domain, Patient Rights, and it is stipulated that: “*Waiting times and queues are managed to improve patient satisfaction and care*.”

More recently, the National Department of Health has established the ‘Ideal Clinic’ programme as a way to address the deficiencies in primary health care clinics in the public sector [17]. The Ideal Clinic Manual identifies 10 components, and 32 sub-components that need to be tracked in order to realise an Ideal Clinic – of these patient waiting time is a core commitment.

A number of studies have been conducted to identify bottlenecks in patient flow and the main factors leading to long wait times in clinics across Africa [4-6]. However, causes of long wait times seem to vary between clinics. While national guidelines have been created to improve delivery models of health care through implementing fast queue systems, increasing access to services on weekends and after-hours, and developing benchmarks against which health care facilities may be assessed in minimizing waiting times [16, 18], local facilities generally set their own norms and guidelines [19].

Literature has shown that appointment scheduling lies at the intersection of efficiency and timely access to health care services [20]. In South Africa, most public health care facilities rely on a system of booking patient appointments scheduled by day, and not by set time (personal communication, July 22, 2015). Operation Phakisa’s Ideal Clinic Realisation and Maintenance Lab identified a number of deficiencies with the current booking system, namely, that (i) patients are provided with dates, and no appointment time with the result that patients arrive early in the morning and create an early-morning bottleneck; (ii) There is no reminder system and patients often miss their appointments; (iii) patients are not consulted on a date that would best suit them, with the result that patients do not adhere to their appointment dates as they are not convenient [21].

This suggests that a poor appointment system may exacerbate long waiting times and impact patient satisfaction and health outcomes. To mitigate these problems, many providers have been advised to create standards around patient wait times; yet, discrepancy often exists between patient and provider norms. Adhering to fixed hours and introducing appointment systems that encourage patients to spread their arrival times throughout the day have been recommended, as well as flexible work shifts that cater to employed patients based on their preferences [9, 19]. In this study, we aimed to explore the benefits and challenges associated with the current system of booking appointments and the feasibility of an alternative booking system from the perspective of both patients and health care providers at a dedicated HIV treatment outpatient clinic set within a large, secondary, public hospital in Johannesburg, South Africa.

## Methods

### Study design

We conducted a cross-sectional, descriptive study. From April and August 2014, trained interviewers conducted surveys with patients and semi-structured interviews with health care providers. The interviews with patients were meant to assess their perspectives on the current appointment system as well as hypothetical, alternative systems. The interviews with providers were meant to assess the reasons for the current appointment system, its strengths and weaknesses, and the potential for alternative appointment systems. We also monitored the total length of the patients clinic visit, their movement through the clinic and time spent with health care workers in order to identify potential bottlenecks. Finally, we linked the medical records of enrolled patients to their interviews and their visit length to determine whether the health and treatment of the patient influenced their appointment preferences.

### Study setting

The study was conducted in Johannesburg, South Africa at a public sector, outpatient HIV treatment clinic set within a large, urban secondary-level teaching hospital. The clinic was established in 2004 and functions as a teaching facility for community service doctors and registrars using a modest clinical staff [22, 23]. According to the sites programmatic data the clinic conducted over 42,000 medical visits and over 61,000 pharmacy visits in 2014. The clinic is open to patients Monday through Thursday from 7:30am-3:30pm, with patient visits starting at 8:00am. Clinic appointments are scheduled by date and not by a set time or time range.

### Study population

For the patient interviews we recruited from the queue of patients waiting for medical appointments at the facility, that is those patients presenting for a pharmacy only visit (i.e. drug pickup) were not eligible. During the study period all patients were required to have a quarterly (every 3 months) routine medical visit where they saw a nurse and/or doctor. In addition to that any patient that was either identified as having or reported a medical problem was also required to have a medical visit. Patients were eligible for the study if they were an adult (≥ 18 years old) patient who had been on ART for at least six months at the study facility and intended to continue their treatment at the study facility. Patients were excluded if they were receiving ART for the prevention of mother-to-child transmission of HIV or not on life-long ART.

We invited all eligible health care workers (HCW) at the site to participate. All adult (≥ 18 years old) HCWs were eligible if they provided HIV care and / or treatment at the study site, and had done so for at least six months if not in a management position.

All participants – both patients and health care providers – provided written informed consent prior to being interviewed. The patient recruitment process is summarised in Figure 1 below. A total of 245 patients were enrolled.

**Figure 1:**
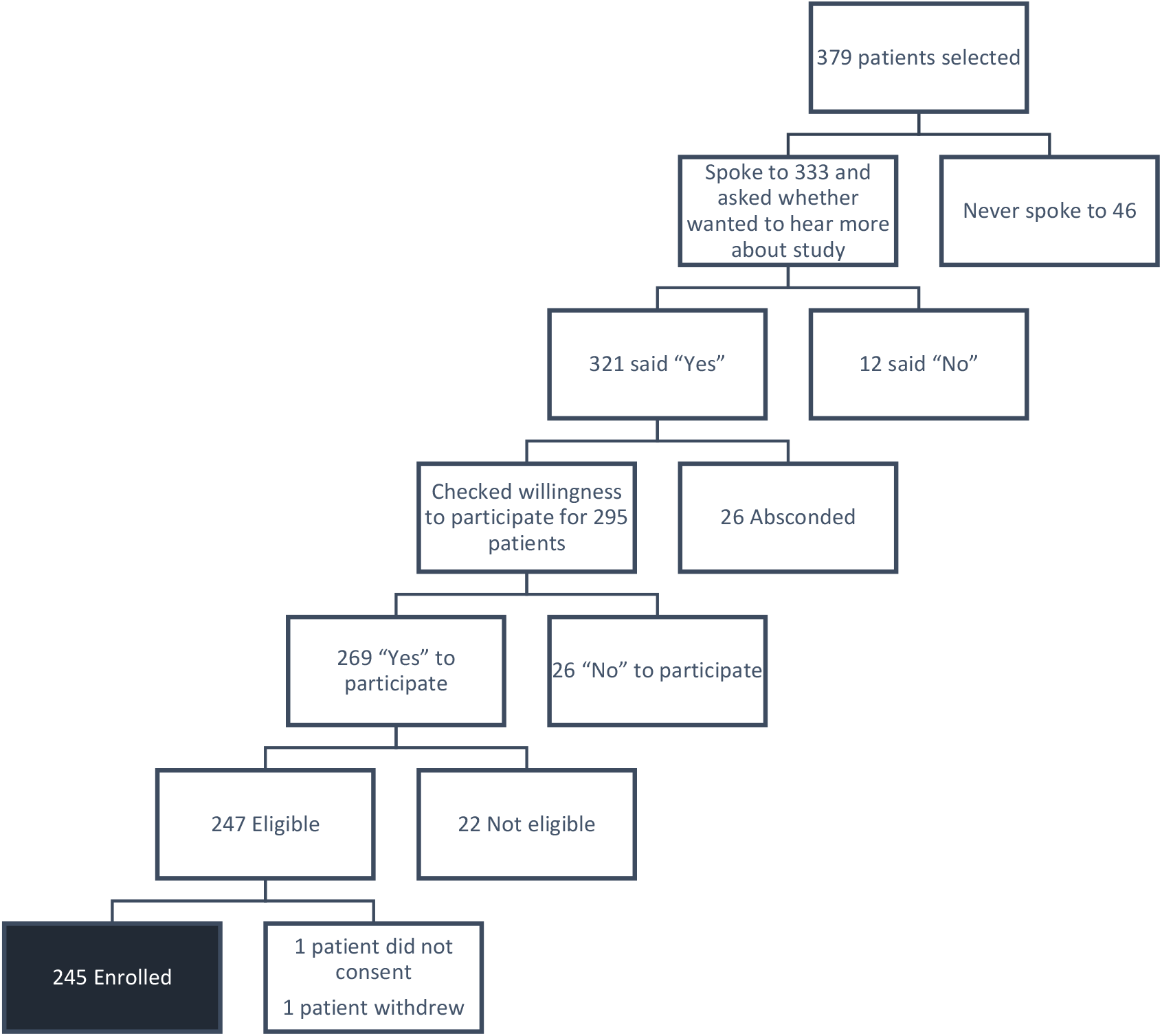
Recruitment Flow chart.

**Figure 2:**
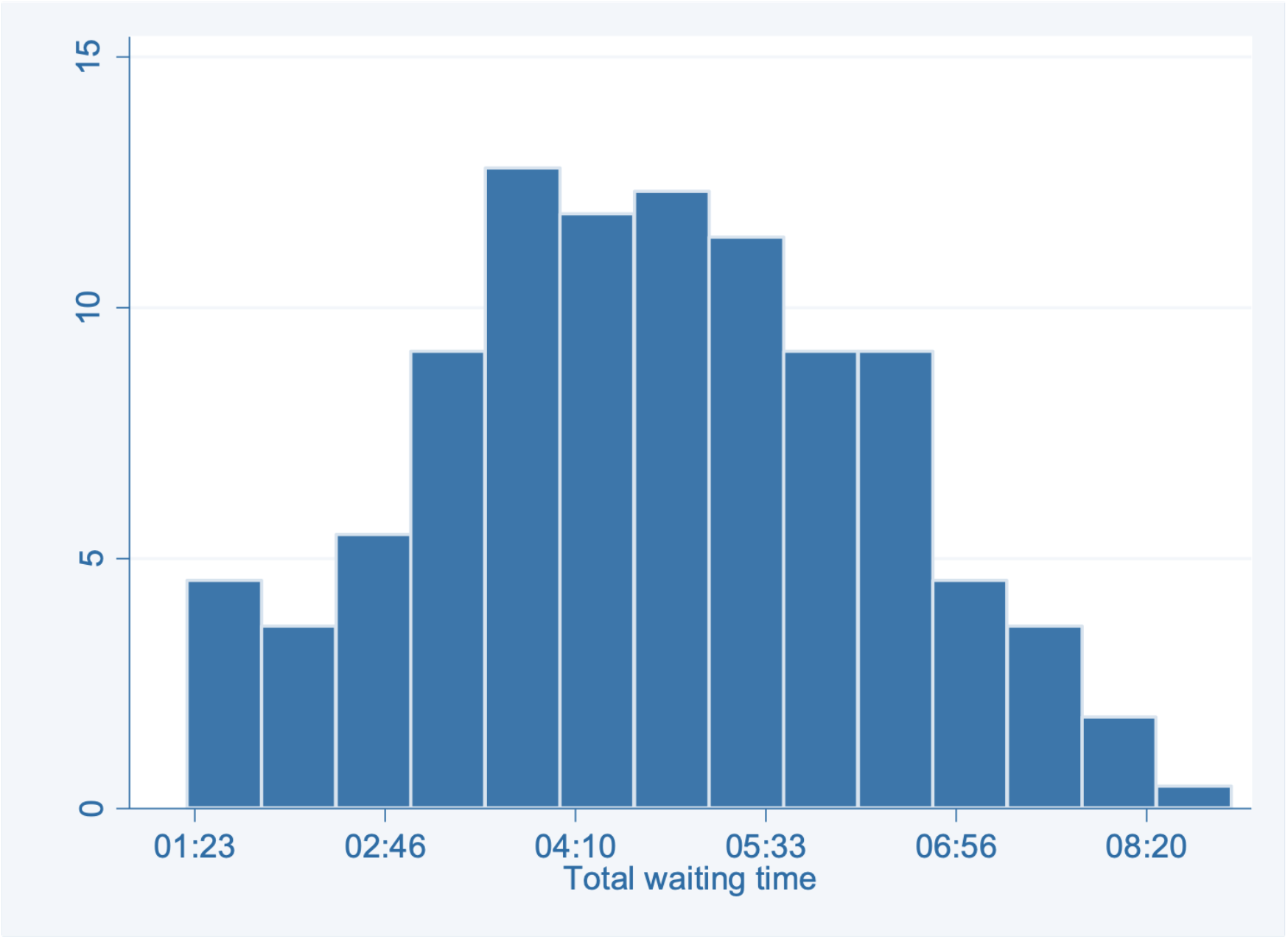
Distribution of patients’ total waiting time.

**Figure 3:**
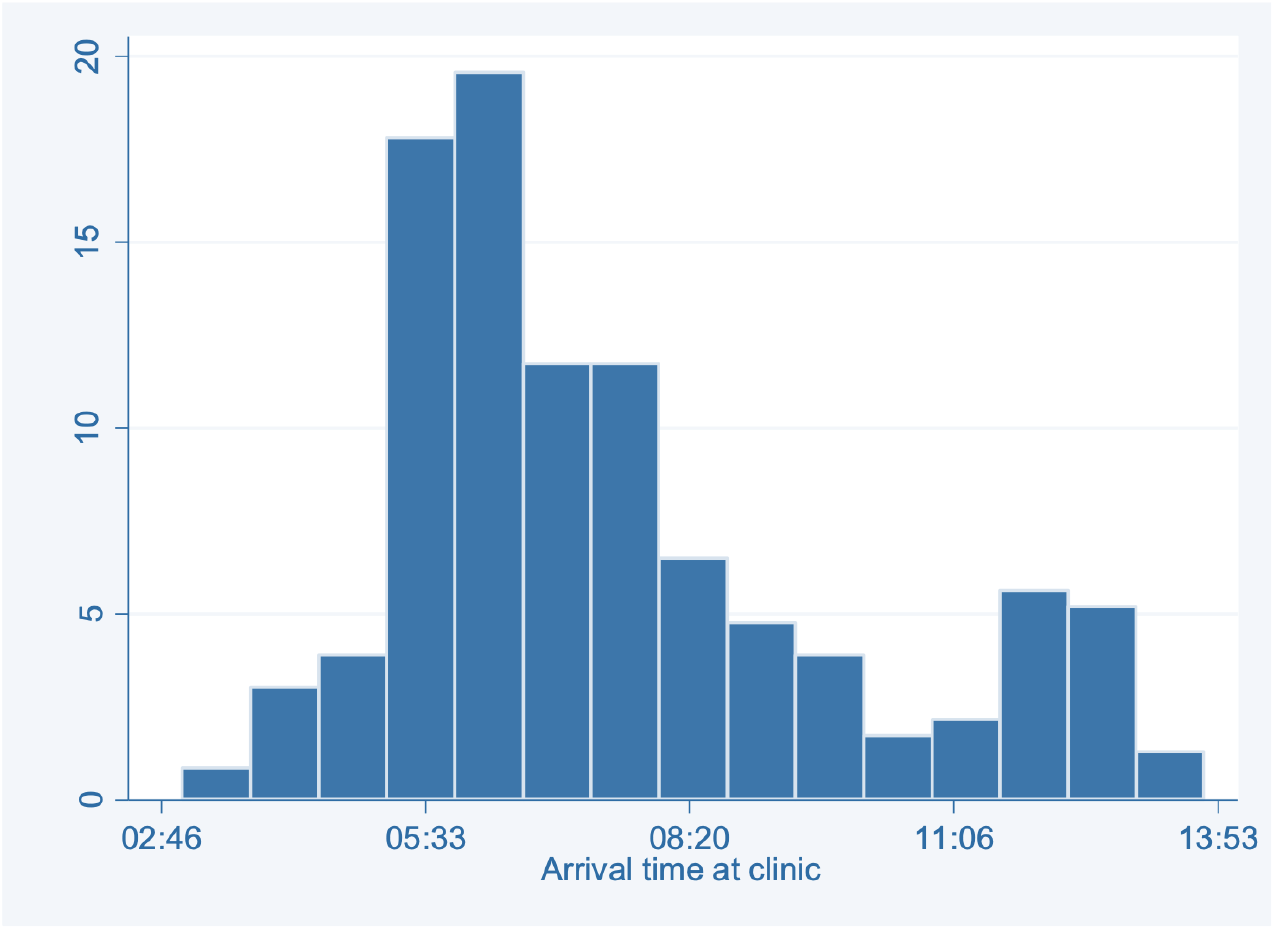
Distribution of patients’ arrival time at the clinic.

### Study procedures and data analysis

We selected patients to approach for recruitment using a systematic sample with a random starting point generated in advance using SAS (version 9.4). The size of the sampling interval was based on average, daily patient volume as estimated using clinic data. We also determined a desired sample size in advance of recruiting patients of 246. We conducted face-to-face interviews with providers and patients using two different standardized questionnaires. Provider interviews were conducted in English; whereas interviews with patients were conducted in English, Zulu or Sotho.

The patient interview included some financial questions. For example, we asked about their average monthly income, costs of travel to/from the facility, etc. This data was collected in South African Rands (ZAR) and is presented here in 2014 US dollars based on an average exchange rate for the study period^1^ and inflation rates for South Africa as published by the Reserve Bank of South Africa.

We also observed the activities of participating patients and requested that clinic staff indicate on a study card the start and stop time of their interactions with the patients as the patients moved through the clinic. To assess the time required for the visit as a whole, the study staff asked the patients what time they arrived at the clinic during the interview. The staff also observed and recorded the patients’ departure times from the clinic.

Finally, we reviewed the patients’ electronic medical records as maintained at the study site. This included collection of the following: date of first HIV-positive diagnosis, most recent CD4 count and date, most recent viral load and date, current ART regimen and date initiated, any missed ART appointments in the past six months, presence of opportunistic infections or other HIV/AIDS-related symptoms in the last six months and any other health concerns/conditions reported or diagnosed in the last six months.

We conducted all quantitative analyses using STATA software (version 14). We generated descriptive statistics to present patient characteristics and participants’ experiences and perspectives on the current and hypothetical, alternative booking systems. We present categorical data as frequencies and proportions; while continuous data is presented as medians with interquartile ranges (IQRs). Open-ended questions were collaboratively analyzed in Excel using a thematic analysis approach. For observational visit timing data, we again used STATA to calculate the average waiting times per patient and per activity.

Approval to conduct the study was obtained from the study site and the Human Ethics Research Committee at the University of the Witwatersrand.

## Results

### Participant characteristics

As shown in Figure 1, we approached 333 individuals during recruitment of patients at the clinic. Of those, we enrolled 245 patients (158 (64.5%) female and 87 (35.5%) male) (Table 1). The median age for all participants was 41 years (IQR 36-47). The majority (234 (95.5%)) of the enrolled patients identified as Black. Roughly half of all patients were married or cohabiting (133, 54.3%), and most (213, 88.4%) had at least one child.

**Table 1:**
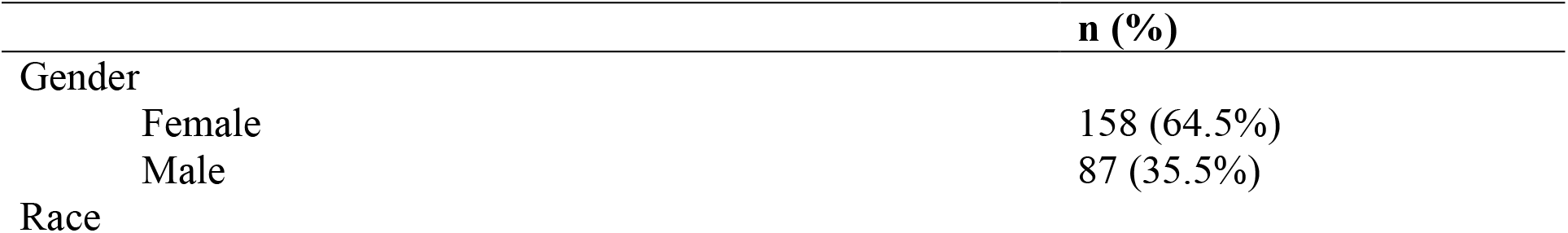
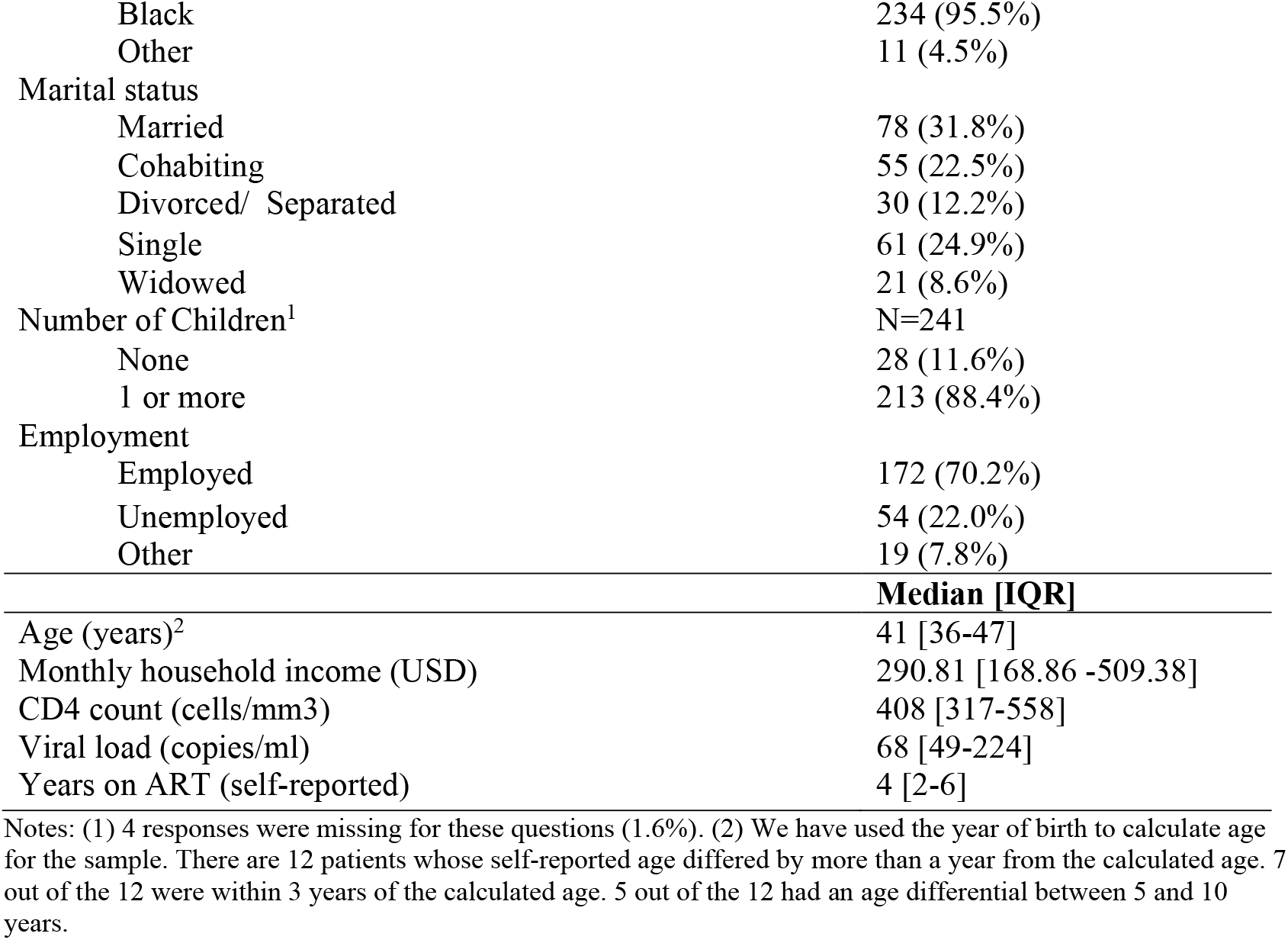
Baseline characteristics for patients at HIV-clinic (N=245)

|  | n (%) |
| --- | --- |
| Gender |  |
| Female | 158 (64.5%) |
| Male | 87 (35.5%) |
| Race |  |
| Black | 234 (95.5%) |
| Other | 11 (4.5%) |
| Marital status |  |
| Married | 78 (31.8%) |
| Cohabiting | 55 (22.5%) |
| Divorced/ Separated | 30 (12.2%) |
| Single | 61 (24.9%) |
| Widowed | 21 (8.6%) |
| Number of Children<sup>1</sup> | N=241 |
| None | 28 (11.6%) |
| 1 or more | 213 (88.4%) |
| Employment |  |
| Employed | 172 (70.2%) |
| Unemployed | 54 (22.0%) |
| Other | 19 (7.8%) |
|  | **Median [IQR]** |
| Age (years)<sup>2</sup> | 41 [36-47] |
| Monthly household income (USD) | 290.81 [168.86 -509.38] |
| CD4 count (cells/mm3) | 408 [317-558] |
| Viral load (copies/ml) | 68 [49-224] |
| Years on ART (self-reported) | 4 [2-6] |

Notes: (1) 4 responses were missing for these questions (1.6%). (2) We have used the year of birth to calculate age for the sample. There are 12 patients whose self-reported age differed by more than a year from the calculated age. 7 out of the 12 were within 3 years of the calculated age. 5 out of the 12 had an age differential between 5 and 10 years.

Considering the patients’ economic means, 172 (70.2%) reported being employed (including formal, informal, and self-employment). They also reported a median monthly household income of USD 291. The patients had been taking ART for a median of four years. The median CD4 count for all patients was 408 cells/ul, and the median viral load was 68 copies/mm^3^.

During the study, the clinic employed 34 health care providers. We approached 23 and six were interviewed. This included one medical officer, two professional nurses, two counsellors, and one pharmacist. Three of these individuals were employed in a management position at the time of the interview. The average length of time working at the facility was 5 years (range 2 - 8 years).

### Accessing services

Patients traveled a median of 45 minutes to the clinic, with a median arrival time at 6:50 am (Table 2). Self-reported waiting time at the clinic was a median of five hours. This corresponded closely with the actual median waiting time, based on our observations and calculations, of 4 hours 43 minutes, which ranged from 1 hour 29 minutes to 8 hours 57 minutes.

**Table 2:** Patients’ reported burdens and costs related to medical appointments (n=245)

|  | **Median [IQR]**<br>**n (%)** |
| --- | --- |
| Length of travel (minutes) to clinic | 0:45 [0:30-01:00] |
| Arrival time at clinic | 6:50am [5:45am-8:29am] |
| Self-reported waiting time (hours) at clinic | 5:00 [4:00-6:00] |
| Calculated total waiting time (hours) at clinic | 4:43 [3:38-5:52] |
| No. of patients who lost income to come to clinic (N=245) | 47 (19.2%) |
| Mean income lost per clinic visit<sup>1</sup> (N=44) | USD 31.89 |
| No. of patients using public transport to clinic | 191 (78%) |
| Mean cost of traveling to clinic from house (one way) (N=215) | USD 2.44 |
| No. of patients who visit current or other healthcare facility for other health-related issues (non-ART) (N=245) | 45 (18.4%) |
| Frequency with which patients visit another facility for other most accessed health service (N=45) |  |
| At least every two months | 28 (62.2%) |
| Quarterly to annually | 15 (33.3%) |
| Frequency with which patients normally attend study clinic<sup>2</sup> (N=245) |  |
| At least every two months | 79 (32.2%) |
| Quarterly and six-monthly | 156 (63.7%) |

**Notes:** (1) 44 out of the 47 patients who reported losing income to come to the clinic gave an estimate of the income lost. 3 responses were missing. (2) There were 10 patients who did not fall into these categories stating that visit frequencies varied and depended on their state of health.

Nearly 20 percent of patients reported losing income to attend each appointment, losing an average of USD 32 per visit. Patients reported an additional average transport cost of USD 2.44 to get to the clinic one way. Nearly 20% of patients also cited that they must make a separate appointment for their other health-related issues. Of these patients, 62% must visit another healthcare facility at least every two months for these issues. Sixty-four percent of patients reported only needing to attend the study clinic every three to six months.

The figure below shows the large variation in patients’ total waiting time where the vast majority of patients wait more than three hours (86%), with some waiting almost 9 hours, and others just over an hour^2^.

We would expect patients’ arrival time to be more evenly dispersed throughout the day. However, some 65% of patients arrive before the clinic opens at 08:00 am. There is another bulge around 12:00 pm in the day. This pattern of arrival time appears to exacerbate total wait time. A significantly negative relationship between arrival time at the clinic and the total time that the patient spends at the clinic is displayed Figure 4.

**Figure 4:**
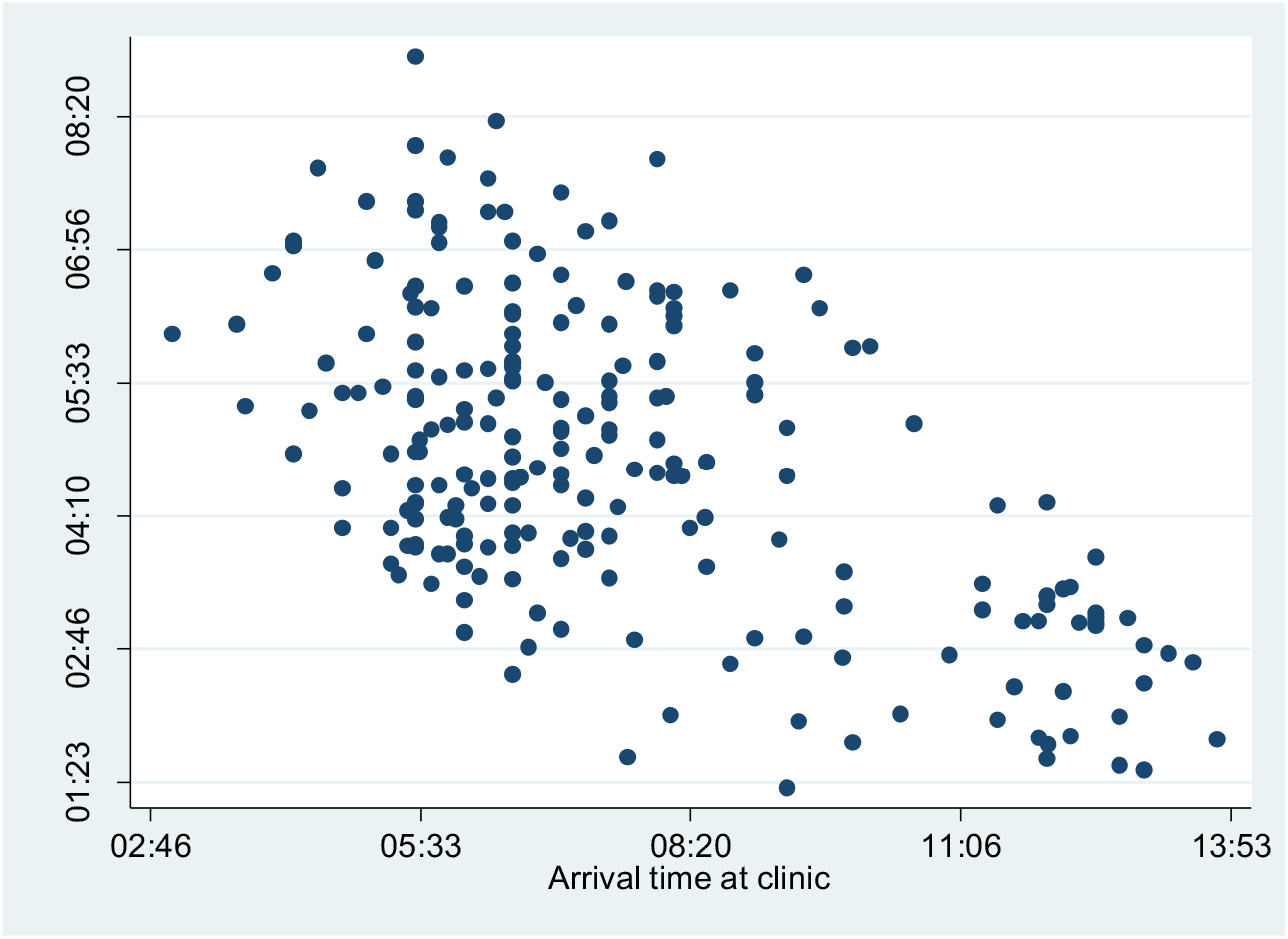
Scatter plot of patients arrival time and total patient waiting time.

### Drivers of arrival time

When patients were asked about their choice of arrival time, most (62.5%) reported wanting to be at the head of the queue, presumably so that they could be seen first and could leave early (see Table 3). Over 13% of patients’ arrival time was dictated by when they could find transport to the clinic, and 12.4% chose their arrival time based on their work schedule. Fourteen percent were not originally scheduled for an appointment on the day they arrived. Of those arriving earlier or later than their original appointment, 45.5% did so because they could not miss work. Thirty-nine percent of all patients reported ever having missed a scheduled appointment. The reason most cited for missing an appointment was once again due to work (39.6%).

**Table 3:**
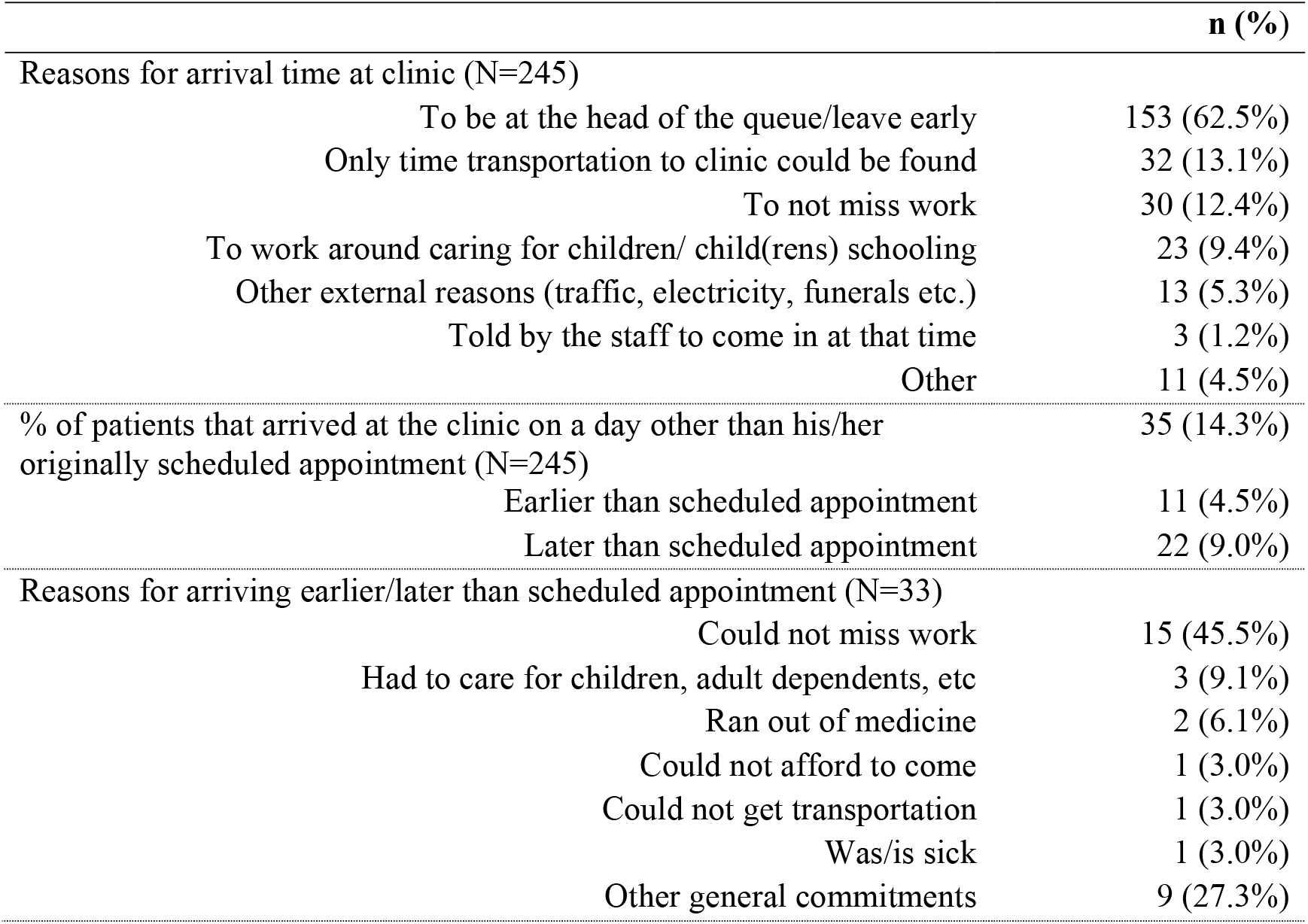
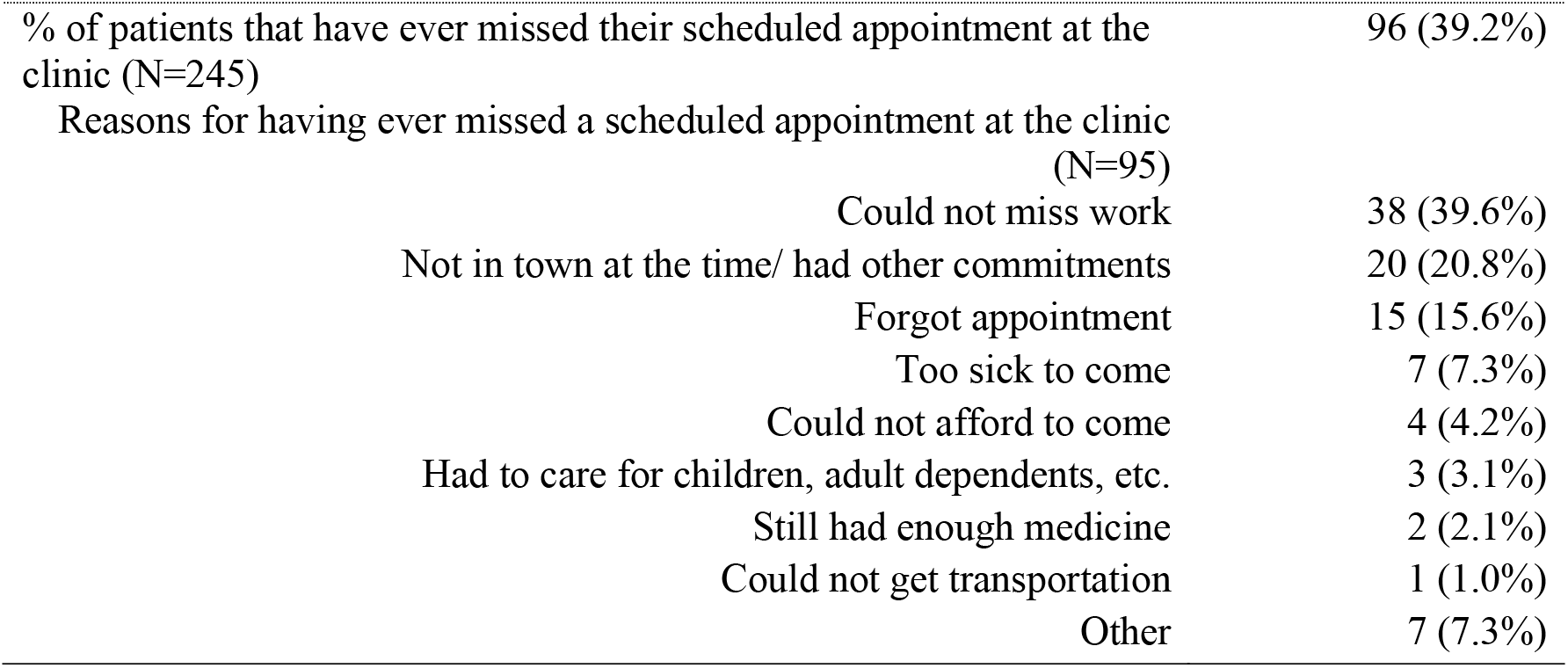
Drivers of arrival times at the clinic (N=245)

|  | n (%) |
| --- | --- |
| Reasons for arrival time at clinic (N=245) |  |
| To be at the head of the queue/leave early | 153 (62.5%) |
| Only time transportation to clinic could be found | 32 (13.1%) |
| To not miss work | 30 (12.4%) |
| To work around caring for children/ child(rens) schooling | 23 (9.4%) |
| Other external reasons (traffic, electricity, funerals etc.) | 13 (5.3%) |
| Told by the staff to come in at that time | 3 (1.2%) |
| Other | 11 (4.5%) |
| % of patients that arrived at the clinic on a day other than his/her<br>originally scheduled appointment (N=245) | 35 (14.3%) |
| Earlier than scheduled appointment | 11 (4.5%) |
| Later than scheduled appointment | 22 (9.0%) |
| Reasons for arriving earlier/later than scheduled appointment (N=33) |  |
| Could not miss work | 15 (45.5%) |
| Had to care for children, adult dependents, etc | 3 (9.1%) |
| Ran out of medicine | 2 (6.1%) |
| Could not afford to come | 1 (3.0%) |
| Could not get transportation | 1 (3.0%) |
| Was/is sick | 1 (3.0%) |
| Other general commitments | 9 (27.3%) |
| % of patients that have ever missed their scheduled appointment at the clinic (N=245) | 96 (39.2%) |
| Reasons for having ever missed a scheduled appointment at the clinic (N=95) |  |
| Could not miss work | 38 (39.6%) |
| Not in town at the time/ had other commitments | 20 (20.8%) |
| Forgot appointment | 15 (15.6%) |
| Too sick to come | 7 (7.3%) |
| Could not afford to come | 4 (4.2%) |
| Had to care for children, adult dependents, etc. | 3 (3.1%) |
| Still had enough medicine | 2 (2.1%) |
| Could not get transportation | 1 (1.0%) |
| Other | 7 (7.3%) |

### Current service provision – Challenges and costs with the current system

According to most of the health care providers, patient appointments are booked based on the amount of medicine dispensed to the patient by the pharmacist. All but one of the patients (99.6%) reported booking their most recent appointment with the booking clerk before they left the clinic at their last visit suggesting that patients can only receive appointments if they are physically present at the clinic.

Health care providers believed that the largest challenge for patients with the existing system of booking appointments is that it is not flexible enough for patient work schedules. Stigma was also reported by some providers as a barrier that drives patients to resist being down referred to their own local clinics and instead attend the current clinic, while issues regarding transport or finding money for transport were also cited by some providers as the largest challenge with the current system.

For the providers themselves, half believed that there is not enough staff to support the current appointment system, and in particular, that doctors rotate too often. One provider suggested the reason for this being that doctors are not adequately incentivized. Half of the providers also cited poor staff attitudes as an obstruction to current appointment system, and some report that the inconsistency in the flow of patient volume across days makes it hard to effectively plan their work days. One provider mentioned that patients simply do not attend their appointments when they are told to; a comment that is reiterated throughout the provider interviews.

When patients were asked about the challenges they face with the current appointment system, they most often cited overcrowding and long queues (29%). Nearly 5% reported that it was not flexible enough with their work schedules. However, most (65%) said there were no challenges with the current system (Table 4).

**Table 4:** Patient challenges and benefits of current system of booking appointments at the clinic (N=245)

|  |  | n (%) |
| --- | --- | --- |
| % of patients who cited the following challenges with the current booking system |  |  |
|  | Creates overcrowding and long queues | 70 (28.6%) |
|  | Not flexible with work schedule | 12 (4.9%) |
|  | May not be enough available medical resources due to overcrowding | 1 (0.4%) |
|  | No challenges with this kind of system | 159 (64.9%) |
| % of patients who cited the following benefits with current booking system |  |  |
|  | Showing up at a time that is most convenient | 121 (49.4%) |
|  | Working outside operational clinic hours | 4 (1.6%) |
|  | Easy to find care for children, adults dependents, and family | 2 (0.8%) |
|  | Easy to find transportation to the clinic | 6 (2.5%) |
|  | No benefits with this kind of system | 72 (29.4%) |

### Current service provision – Benefits with the current system

Despite the challenges reported by the providers, they believed the current system provides a number of benefits. All but one of the providers believed that the largest patient benefit of the current system is that patients can show up at their own convenience, and thereby work around their own schedules. Some mentioned that the appointment schedule is such that it creates social groups for patients that routinely attend the clinic together. The notion of ‘convenience’ is also reported as a benefit to providers; half believed the current system allows them to control their workload more effectively.

Half also believed the current system is “very effective” in ensuring patients receive the medical services they need in a timely manner, however providers expressed concerns such as patients not complying with their scheduled visit, issues among staff with time management and providers arriving at the clinic late that may cause bottlenecks, and faulty SMS appointment reminders as reasons that may affect the timeliness of health care provision. Half of providers also feel like the clinic does not have enough staff to service the patients that come through the clinic per day. Some reasons cited for this are the generally large volume of patients, the shortage of doctors, and the inefficient use of staff time.

When patients were asked about the benefits they face with the current appointment system, they most often cited showing up at their own convenience (49%). Twenty-nine percent did not believe that there are any benefits to the current system (see Table 4).

### Current waiting time and perceptions of service quality

When providers were asked which aspects of delivering health care services to patients were most highly regarded as important, values and attitudes of the staff, that the staff are knowledgeable and well trained, and that medicine is available were most highly rated. Waiting times for the patient were valued lowest on average. When patients were asked about how important the same aspects of healthcare delivery were in terms of their satisfaction with the services they receive, they reported a similar direction of results (see **Table 5**).

**Table 5:** Aspects of health care provision that are important in terms of service quality rated by patients and providers on a scale from 1 [least important] to 10 [most important].

|  | Patients | Providers |
| --- | --- | --- |
| Values and attitudes of staff | 8.4 | 10.0 |
| Staff are knowledgeable and well trained (clinically) | 8.6 | 10.0 |
| Cleanliness of the facility | 8.3 | 9.7 |
| Waiting times at the facility | 5.9 | 9.3 |
| Availability of medicines | 9.7 | 10.0 |

### Provider and patient satisfaction with overall services and waiting times

Most of the providers did not feel that patients wait too long to receive medical services at the clinic, and half said that waiting times have gotten shorter over time. Half of those who feel that waiting times are not long mentioned down referrals as the reason for this. The two providers that felt that patients wait too long believe that recurring issues with staff can cause delays and bottlenecks, issues such as arriving to the clinic too late, taking too many meetings, or bad attitudes/work ethics. All but one of the providers also believed that the patients are satisfied with the amount of time they wait at the clinic, and most think the reason for this is that the appointment system generally runs fine and is quick. However, a couple still acknowledged that there are some delays and bottlenecks at certain stations, which may impact patient satisfaction. Whilst another provider had a negative perception of patients stating that they like to complain for no reason.

When patients were asked about their satisfaction with the health care services they receive, most were satisfied (82.4%) (see Table 6). Significantly less reported being satisfied with the amount of time they wait for services at the clinic (35.6%). When clinic times for the interviewed patients were observed from the time, they arrived at the clinic to the time they left, they stayed an average of 4 hours and 44 minutes at the clinic.

**Table 6:** Patient satisfaction with health care services and waiting times at the clinic (N=245)

|  | Overall services<br>n (%) | Waiting times<br>n (%) |
| --- | --- | --- |
| Not sure | 1 (0.4%) | 4 (1.6%) |
| Very dissatisfied | 8 (3.3%) | 101 (41.2%) |
| Somewhat dissatisfied | 5 (2.0%) | 6 (2.5%) |
| Neutral | 29 (11.8%) | 47 (19.2%) |
| Somewhat satisfied | 26 (10.6%) | 19 (7.8%) |
| Very satisfied | 176 (71.8%) | 68 (27.8%) |

### Alternative scenarios for service provision

In addition to their perceptions on the current system, patients and providers were asked about their opinions on three alternative booking systems involving a more structured appointment schedule: 1) appointments scheduled after traditional work hours or on the weekends; 2) appointments scheduled for a specified time during the day or 3) appointments scheduled for an open morning block or an open afternoon block (i.e., you could arrive at any time during the morning (8h00-12h00) or afternoon (12h00-3h30), but you would have to come in when you were told – either in the morning or the afternoon).

In general, most of the providers believed that the clinic and the patients that attend could benefit from changes to the booking system. All but two wanted to see after hours appointments introduced at the clinic, and all providers strongly believed that the greatest benefit of introducing after hours appointments is the flexibility it gives patients over their work schedules. Providers expressed some other benefits to providing extended hours as well, including job creation to fill after hours posts, freeing up their time for more or different clinical work, and allowing them to control their workloads more conveniently. Half of the providers believed there will be no direct benefit to the staff and that it would not change their job responsibilities.

When asked about the challenges that may arise from offering extended hours, the most expressed concern among providers is that there may not be enough staff to support this type of system. One provider stated that patients will still not show up to their appointments, even if offered extended hours, and another provider believed there may be problems around transport and safety if staff and patients are at the clinic after traditional work hours. All six providers believed that offering appointments during extended hours will decrease waiting times.

Providers were also asked about their opinions on offering patients specified times for their appointments. All but two were interested. Most believed that this type of system would better help in controlling their workloads and would make their work easier, and some thought it would also help control the flow of patients throughout the clinic. All but one believed their job responsibilities would not change with this system and that it would decrease waiting times. However, providers also expressed apprehension with specified appointment times, including concerns over patients not complying with their scheduled times, and some contrarily believed this system would make it harder to control the flow of patients through the clinic.

Providers conveyed the least interest in a system in which appointments are offered in the morning or the afternoon. Half reported that they would like to see this system. Some believed that transport challenges may still exist with this block system, and once again concerns over patients complying with their appointment times arose during some of the provider interviews. Yet, half thought that block appointments would allow them to control their workloads better and some providers thought this system may be more flexible for patients’ work schedules. All but one did not believe this system would change their job responsibilities, and all believed that it would decrease waiting times for patients.

When patients were asked about their opinions on alternative appointment systems, the largest percentage were interested in morning and afternoon appointments. Eighty-eight percent of patients would like to see appointments offered as a block of time in the morning or afternoon, and 85% would like to be offered appointments after work hours. Only 58% were interested in booking appointments at a specific time (see Table 7). Most respondents (79%) believe the largest benefit to morning/afternoon block appointments would be the ability to show up at their own convenience during the block of time.

**Table 7:** Perceptions on different systems of booking a patient appointment (N=245)

| **Perceptions on different booking systems** | **Current system** | **After work hours** | **Specific time** | **Morning/ Afternoon** |
| --- | --- | --- | --- | --- |
| % of patients who would like to see an alternative booking system |  | 208 (84.9%) | 141 (57.6%) | 215 (87.8%) |
| **% of patients who cited the following challenges with each booking system** |  |  |  |  |
| Creates overcrowding and long queues | 70 (28.6%) | 7 (2.9%) | 19 (7.8%) | 10 (4.1%) |
| Not flexible with work schedule | 12 (4.9%) | 9 (3.7%) | 26 (10.6%) | 10 (4.1%) |
| Hard to find care for children, adult dependents, or family | 0 (0.0%) | 6 (2.5%) | 3 (1.2%) | 1 (0.4%) |
| Hard to find transport with these hours | 0 (0.0%) | 20 (8.2%) | 92 (37.6%) | 12 (4.9%) |
| Being turned away if the queues are too long and the clinic closes before medical services are received | 0 (0.0%) | 1 (0.4%) | 2 (0.8%) | 1 (0.4%) |
| Not be enough available medical resources due to overcrowding | 1 (0.4%) | 7 (2.9%) | 3 (1.2%) | 0 (0.0%) |
| No challenges with this kind of system | 159 (64.9%) | 183 (74.7%) | 121 (49.4%) | 210 (85.7%) |
| **% of patients who cited the following benefits with each booking system** |  |  |  |  |
| Showing up at a time that is most convenient | 121 (49.4%) | 172 (70.2%) | 103 (42.0%) | 194 (79.2%) |
| Working outside operational clinic hours | 4 (1.6%) | 80 (32.7%) | 12 (4.9%) | 17 (6.9%) |
| Easy to find care for children, adults dependents, and family | 2 (0.8%) | 10 (4.1%) | 1 (0.4%) | 3 (1.2%) |
| Easy to find transportation to the clinic | 6 (2.5%) | 4 (1.6%) | 0 (0.0%) | 6 (2.5%) |
| No benefits with this kind of system | 72 (29.4%) | 38 (15.5%) | 114 (46.5%) | 33 (13.5%) |

## Discussion

Waiting time satisfaction is an important component of overall patient satisfaction. The targets established by the Ideal Clinic programme define a **3 hour maximum total waiting time** for patients in all clinics; a target of 80% of patients reporting a positive experience of care; and a target of 90% of patients indicating that they are satisfied with their waiting time at the clinic[17]. Whilst patients in this study do not view waiting time as very important (it scored a 5.9/10 where 10 is most important), nor do they see a problem with the current booking system (65% saw no problem), they were not satisfied with waiting times. Only 35.6% of patients indicated that they were somewhat satisfied or very satisfied with waiting times and 86% of patients waited longer than the stipulated 3 hours. Conversely, over 80% were satisfied with the overall service they received suggesting waiting time is not weighted significantly in an overall evaluation of quality care by patients.

This study has explored the benefits and challenges with the current booking system and preferences for alternative booking systems. The role of an effective booking system is to essentially manage demand and reduce waiting times. The current system does not appear to be working effectively evidenced by the fact that there are long median waiting times, and demand is unevenly distributed throughout the day with spikes at particular points in the day, specifically in the early morning. Sixty five percent of patients arrive at the clinic before it opens, and 62.5% said that their arrival time was driven by their desire to be at the head of the queue. The reason for the last point could be due to a belief that they would be served quicker, not turned away, or they would be able to maximise their day off. This however does not appear to occur in practice shown by the negative relationship between arrival time and total wait time. This high early morning demand could be more effectively managed through a reliable appointment schedule. Other reasons for arrival time, such as work and child care commitments could be resolved through after hour appointments. In addition, the high proportion of patients who miss appointments (40% of patients indicated they had a missed an appointment at some point) could be avoided if patients were consulted in the appointment booking process on a date that best suited their schedules.

Of the 15 breakthrough initiatives recommended by Operation Phakisa for the realisation of the Ideal Clinic programme, the first one related to Waiting Times, and more specifically to improving the booking system by implementing a “*functional appointment system for non-emergency patients*” ^3^[21]. It was proposed that appointments will be handled centrally at each clinic, agreed with patients based on their clinical requirement and given for **specific times**^4^.

The Operation Phakisa recommendation of a booking system that provides patients with an appointment for a specific time in the day was the least preferred option in *this* study. Only 57.6% of patients opted for this option and 37.6% citing difficulties with finding transport as one of the main challenges with this system. In comparison, convenience was cited as an important benefit of both the after-hours booking system (70.2% of patients) and the morning/afternoon booking system (79.2% of patients).

Seventy eight per cent of patients take some form of public transport to the clinic (taxi, train or bus). This affects the ability of patients to arrive at a specific time as public transport is unpredictable outside of the rush hour times in the early morning and late afternoon. 13.1% of patients said their arrival time was driven by when transport was available. As such, the flexibility offered by the morning/afternoon booking system might better correlate with the transport needs of the patients. It might also be a good intermediate step prior to implementing specific time appointments as patients need time to build their trust in the booking system. The 3 hour waiting time target is feasible in a morning/afternoon block session where the potential waiting time is halved to 4 hours. Working adults should only need to miss at most a half day of work.

## Limitations

There have been changes to the clinic over time which may have improved patient experiences and waiting times. These include pharmacy automation, improvements in patient flow processes (patient file retrievals), chronic medication pick up points, and down-referral for stable^5^ patients which have contributed to declining patient volumes. All these factors would have had an effect on patient and providers’ perceptions of services, waiting times, and booking systems. As such, preferences for different booking systems are likely not to remain stable given these changes.

This was an exploratory study, meant to assess the feasibility of booked appointments from the perspective of providers and patients. This study was conducted prior to the Ideal Clinic proposal instituting time-specific appointment booking system. It remains to be seen how the Ideal Clinic booking system will be implemented and whether adjustments will be made. The patient preferences for different booking systems presented in this study are important considerations in designing a new and better booking system and should be incorporated in the Ideal Clinic booking system going forward.

Changing the booking system, as proposed in this study, is possible. Whilst no specific booking system intervention has been tested, when presented with the hypothetical booking alternatives, health care providers did not think there would be obstacles to implementation and that it could be done if it would improve the patient experience. Furthermore, providing booked appointments during existing working hours should be scalable to most facilities, as it requires operational restructuring instead of additional resources. Extending operating hours may be more challenging as it involves shift work and potentially additional staff.

A formal costing of the suggested alternatives has not been done. However, providing booked appointments should not add any additional cost. By spreading the patient load it may allow better utilisation of current resources, including human resources. It could also reduce the cost to the patient of extended waiting hours. Extending operating hours, on the other hand, would likely result in additional costs. Though improved patient retention could result in cost savings for the health system as a whole.

## Conclusion

In this study, we aimed to explore the benefits and challenges associated with the current system of booking appointments and the feasibility of an alternative booking system from the perspective of both patients and health care providers at a dedicated HIV treatment outpatient clinic set within a large, secondary, public hospital in Johannesburg, South Africa.

The results indicate that patients highly value convenience. This may be explained by the need to be flexible around work schedules and transport options. Future research is needed to test the impact of converting to a booking system in which patients are given a morning or afternoon block appointment rather than a day, and to explore the cost implications of after-hours services. Extended working hours would assist patients who lose income as a result of attending the facility during working hours. Both booked morning/afternoon block appointments and after hours services could potentially contribute to improved patient experiences and retention in care in a high-prevalence HIV setting.

## Data Availability

All data produced in the present study are available upon reasonable request to the authors, approval from the Human Research Ethics Committee (University of Witwatersrand) and approval from the study site who owns the clinical data.

## Footnotes

1 The average Dollar/Rand exchange for the period 1 April 2014 to 30 September 2014 was R10.66/USD.

2 A National Department of Health study found that patient’s total waiting time in clinics ranged from two to seven hours with, on average, 76% of patients’ time in the clinic spent waiting. (Lean diagnostics in Ideal Clinic Pilot Sites).

3 The other initiatives to reduce waiting time included in the recommendations are: (1) Establish provincial health call centres to provide advice and reduce unnecessary burden on clinics (2) Set up an SMS-based communication platform to enable the communication of individualised patient information, such as appointment reminders (3) Improve efficiency of patient flow (4) Standardise paper filing processes (5) Support clinics to adjust hours/days of operation (6) Implement an electronic queue management system (7) Communicate clear expectations for waiting times and process of care (8) Evaluate, improve and communicate patient experience of care and waiting times as a Key Performance Area.

4 Other standard criteria for the proposed appointment system included: (1) Each clinic will determine daily targets for each date and time, informed by patients’ conditions and needs as well as staff capacity per day. (2) To balance patient influx and staff workload under the new process, each clinic will have a well-structured, centralised appointment-booking system. (3) Appointments for medication pickup will be given at designated alternate sites where available. (4) Non-booked patients will be seen on the same day but waiting times may be longer as priority will be given to patients with a regular appointment. (5) The appointment system will be linked to the automated patient identification system once this is operational. (6) Patients will receive SMS reminders once an appropriate system has been developed (leveraging experiences like MomConnect).

5 Eligibility to be down-referred includes being on ART for at least 11 months, an undetectable viral load in the previous 10 months, stable weight, CD4 count >200 cells/mm^3^, <5% weight loss over the last three visits and no opportunistic infections.

